# Development of reference ranges of vital signs in UK children: comparison with international centiles

**DOI:** 10.1101/2022.09.11.22279812

**Authors:** Nicky Taylor, Petra A Wark, Jane Coad, Andrew Prayle, Joseph C. Manning

## Abstract

**Background:** Children and Young People (CYP) with acute illness require routine assessment of their physiological observations. There is no agreement as to what the standard reference ranges of vital signs in children and young people are. Existing reference ranges of vital signs that are currently used in clinical practice are minimally supported by empirical evidence. They are also sometimes conflicting.

**Methods:** We conducted a cross-sectional study using 66356 admission episodes to analyze routinely collected age-specific respiratory rate, heart rate, and blood pressure observations of CYP aged 0-19 years old at hospital discharge. Quantile regression with Restricted Cubic Splines was used to model age-specific centiles. These were then compared with standard reference ranges and literature.

**Results:** New centile charts for vital signs are presented. Advanced Paediatric Life Support (APLS, 6th Ed.) reference ranges for respiratory rate and blood pressure poorly aligned to the centiles derived in this study although the centiles for heart rate align well. Variance was also demonstrated between the study centiles and those from the clinical papers, with the greatest differences seen in the upper centiles. Similarly, in comparison with APLS reference ranges, heart rate showed best alignment.

**Conclusions:** This is the first-time physiological observations of CYP in a UK Children’s hospital have been described and centile charts developed. Current widely used reference ranges especially those for Heart Rate and Respiratory Rate are not fit for purpose when evaluating whether the vital signs of a child are normal or otherwise.

## INTRODUCTION

Vital signs are an integral part of a patient’s assessment and pivotal to clinical decision making.^1^ Insight into normal and abnormal ranges is central to assessing the sick child, and for ongoing management of acute patients.^2^ This plays a key role in primary, secondary, and critical care, and forms the basis of many early warning scores.^3,4^

Multiple systematic reviews regarding the vital signs of children and young people (CYP) conclude that there is little empirical evidence to support the reference ranges used in clinical practice.^1,5,6^ There are several conflicting observation reference ranges based on centiles for normal vital signs.^1,3,7^ To date, there is no consensus as to what constitutes ‘normal’ vital signs for paediatric patients.

We used routinely collected vital signs from patients at the point of discharge from hospital to develop age specific centile charts for respiratory rate (RR), heart rate (HR) and blood pressure (BP) in hospitalized CYP. We then compared these to the paediatric reference ranges cited by the Advanced Paediatric Life Support (APLS) Guidelines (UK)^8^ and previously published vital signs reference data. ^3,7,9^

## METHODS

### Study setting and participants

Vital signs data from a tertiary children’s hospital in the United Kingdom that were routinely collected between September 2014 to December 2019 was used for this study. The analysis has not been updated with data from 2020-2021 due to the impact COVID has had on the patient population during this time. The hospital has approximately120 beds across 12 ward areas. The critical care unit and emergency department care for approximately 22,000 inpatient admissions per year, roughly comprising 1.5% of the 1.5 million paediatric admissions in the UK.^10^ We excluded data on patients admitted to Neonatal Intensive Care and Pediatric Critical Care as these CYP may have deranged vital signs. All patients aged 0-19 years from the children’s wards were eligible for this study.

### Ethical considerations

A review by the research ethics committee was not required as data was de-identified by the hospital’s own data analysis team. As only de-identified data were used by the research team, the study was also exempt from the General Data Protection Regulations (GDPR) consent requirements.^11^ Personal identifiable information such as NHS number, name and date of birth were not shared with the research team. The hospitals’ Research and Innovation department along with the Data Guardian granted permission for the research study.

### Data collection

Vital signs were recorded using an electronic observation platform (e-Obs) that was linked automatically to the electronic patient records. Once a patient is admitted to the hospital, the e-Obs system alerts the nurse in charge of the admission (via an iPod device). This alert remains highlighted on the system until the observations are inputted. The hospital’s observation policy mandates a minimum of four hourly observations for 24 hours, then a minimum of once every 12 hours for the duration of inpatient stay. The e-Obs system flags erroneous clinical observations (i.e., a RR of 300). The system also ensures that a complete set of observations are recorded as the system will not allow the observation to be submitted if a mandatory observation is omitted. At the time of the study, BP was not a mandatory discharge observation, and therefore there are fewer observations for this parameter.

All staff who record observations have undertaken standardized training on patient assessment and the recording of physiological observations. Student nurses are under the direct supervision of a registered nurse. There is a hospital policy in place for the assessment and the recording of a patient’s physiological observations.

All ward equipment is centrally procured which ensures standardization of the machinery, its maintenance and the specific training and competency requirements. The hospital’s policy dictates that staff do not use equipment unless formally assessed as competent and their Medical Equipment Service Unit maintains all the medical devices.

### Data extraction

Data on age, sex, and diagnosis at discharge for each patient were also obtained. For each eligible patient, we used the most recent vital signs data that were routinely collected prior to discharge at the study site, between September 2014 to December 2019. We used the final set of observations prior to discharge, as this represents the closest to “normal” for that patient. Even if this is not entirely representative of the patient’s healthy baseline, we reasoned that treating clinician responsible for the patients considered such vital signs data to be compatible with a safe discharge.

### Data analysis

We excluded clinical observations where the temperature was greater than 38°C or less than 35°C, and then developed centile charts from the remainder. Quantile restricted cubic splines regression was used to create centile charts for RR, HR, and systolic BP (SBP). Knots were placed at the 15, 30, 45, 60, 75 and 90th centiles of the data. Data were analyzed using R (version 3.6.2)^12^ and the quantreg (version 5.55)^13^ and rms (version 5.1-4)^14^ packages.

We compared our centile charts to the following internationally recognized reference range and previously published data: (1) Advanced Paediatric Life Support (APLS) guidelines (UK);^8^ (2) centile charts for children’s HR and RR (USA);^3^ (3) reference ranges and centiles for HR and RR in an pediatric emergency department (Australian);^7^ and (4) Distribution of pediatric vital signs in the emergency department (Korea).^9^ The study groups 5^th^ and 95^th^ RR, HR and SBP centiles were chosen as APLS ^8^ describe their upper and lower limits using these two centiles. The study groups 50^th^ centile is used to visualize the median of the distribution to assist in the comparison.

To understand if the discharge observations were influenced by the diagnostic category of the patient, we undertook two sensitivity analyses: (i) an analysis of RR, excluding respiratory diagnoses and (ii) an analysis of HR and SBP, excluding patients with cardiac diagnoses. We used ICD10 ^15^ codes for diagnosis (main reason for admission) to undertake the subgroup analysis.

## RESULTS

The study included 37,783 patients with median age of 4·4 years [IQR: 1·4 to 11·2], and 44.9% female. Observations were recorded electronically between September 2014 to December 2019. Approximately 80% were inpatients less than two days and 60% of the CYP were admitted via an acute/emergency admissions pathway. During this period 67,055 admission episodes were logged, de-identified, and exported for analysis. Some patients had more than one admission during the study period. Of these episodes, 672 were excluded because of high temperature (>38°C), 27 because of low temperature (>35 °C) and three for missing data. This resulted in a final dataset of 66,353 admission episodes.

There were complete data for HR (n = 66,353) and RR (n = 66,353), with approximately 33% (n= 21,474) of participants had their SBP recorded as was not a mandatory discharge observation.

### Centile chart

Centile charts for RR, HR and BP are shown in Figure 1. Heart rate is recorded as beats/minute, respiratory rate as breaths/minute and systolic blood pressure in mmHg. A transparent point represents each patient. The density of the shading represents the distribution of the underlying data; the darker the shading, the higher the density. Black lines are placed on the 5^th^, 10^th^, 25^th^, 50^th^, 75^th^, 90^th^ and 95^th^ centiles of the corresponding vital sign data.

**Figure 1:**
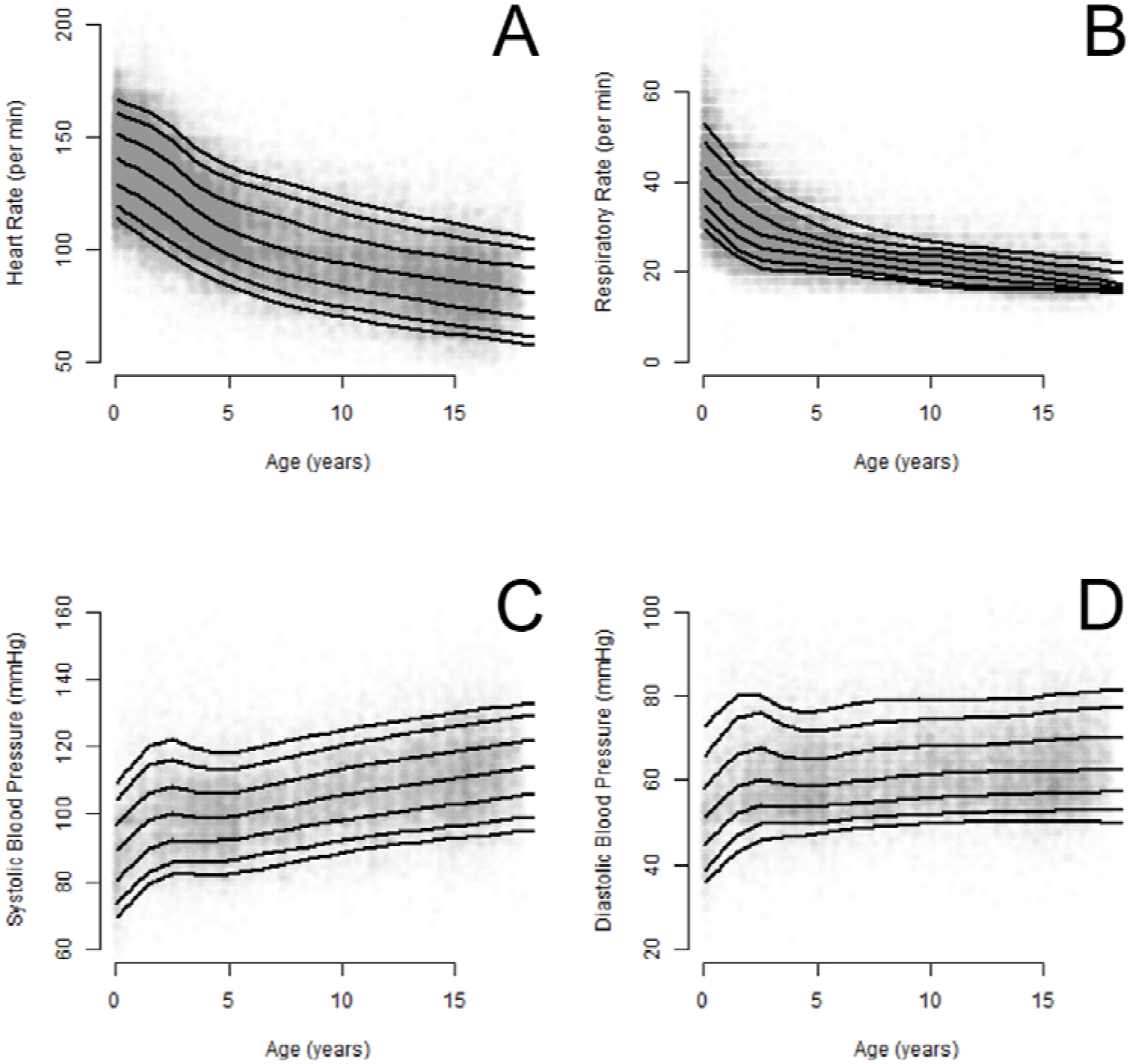
Normal ranges for observations from our cohort for respiratory rate (A), systolic blood pressure (B), heart rate (c) and diastolic blood pressure (D). Each patient is represented by a transparent point. The density of the shading represents the distribution of the underlying data; the darker the shading, the higher the density. The black lines are placed on the 5^th^, 10^th^, 25^th^, 50^th^, 75^th^ 90^th^ and 95^th^ centiles of the corresponding vital sign data.

We compared our centiles to the Advanced Paediatric Life Support (APLS) Guidelines (UK);8 reference ranges (Figure 2). The study group’s respiratory rates are generally higher than those from the APLS reference ranges 8, particularly in the three months to four year age groups. The study groups 95th centile sit above the corresponding centile for APLS, with rates that are 5-10 breaths/minute higher. There is less variation with the zero to three month and over four-year age groups, here the difference between the corresponding centiles is less than five breaths/minute. We also compared our data to reference ranges from previous key reports. ^1,3,7,9^ As we diverged from APLS most for respiratory rate, we present respiratory rate differences (see Figure 3). The respiratory data of Bonafide et al3 show a higher 95th centile compared to both ours and the other published data. The 95th centiles of the other reference ranges are lower than ours, and more in keeping with the APLS reference ranges.^8^ As expected they all have a similar monotonic relationship with age and the greatest changes in respiratory rate occur in the younger aged groups, this is demonstrated in our centiles and those of the other published data. ^3,7,11^

**Figure 2:**
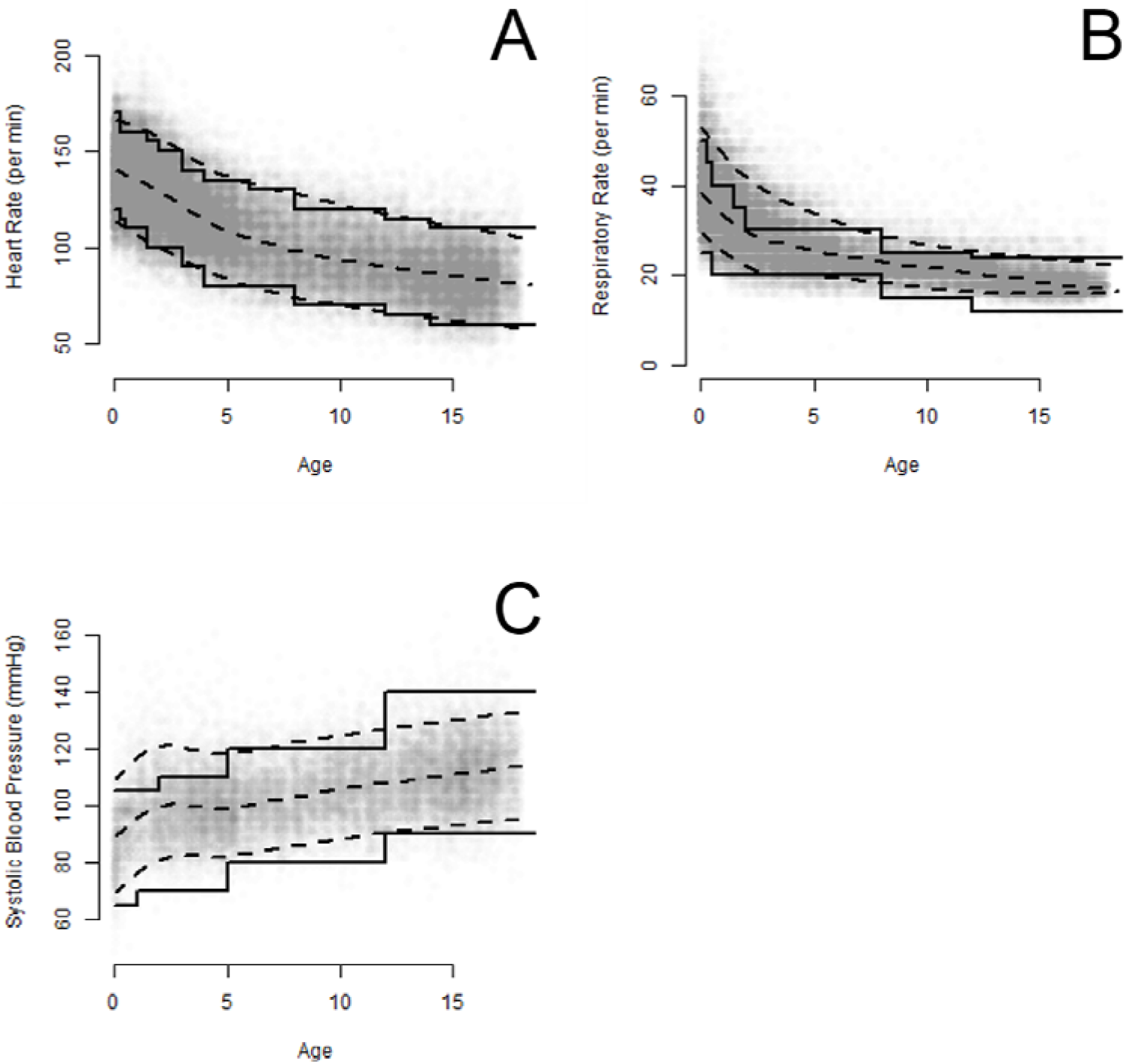
Comparison of the study centile charts with existing reference charts. A-C Comparison between the study cohort data, the centile charts, and Advanced Paediatric Life Support (APLS) observation reference ranges for respiratory rate, heart rate and systolic blood pressure. 5^Th^, 50^th^ and 95^th^ centiles (dotted lines) are compared to APLS reference ranges uninterrupted lines. APLS reports a lower and upper band of blood pressure - for simplicity, we compare the lower end of the lower band to our 5^th^ centile, and the upper end of the upper band to our 95^th^ centile.

**Figure 3:**
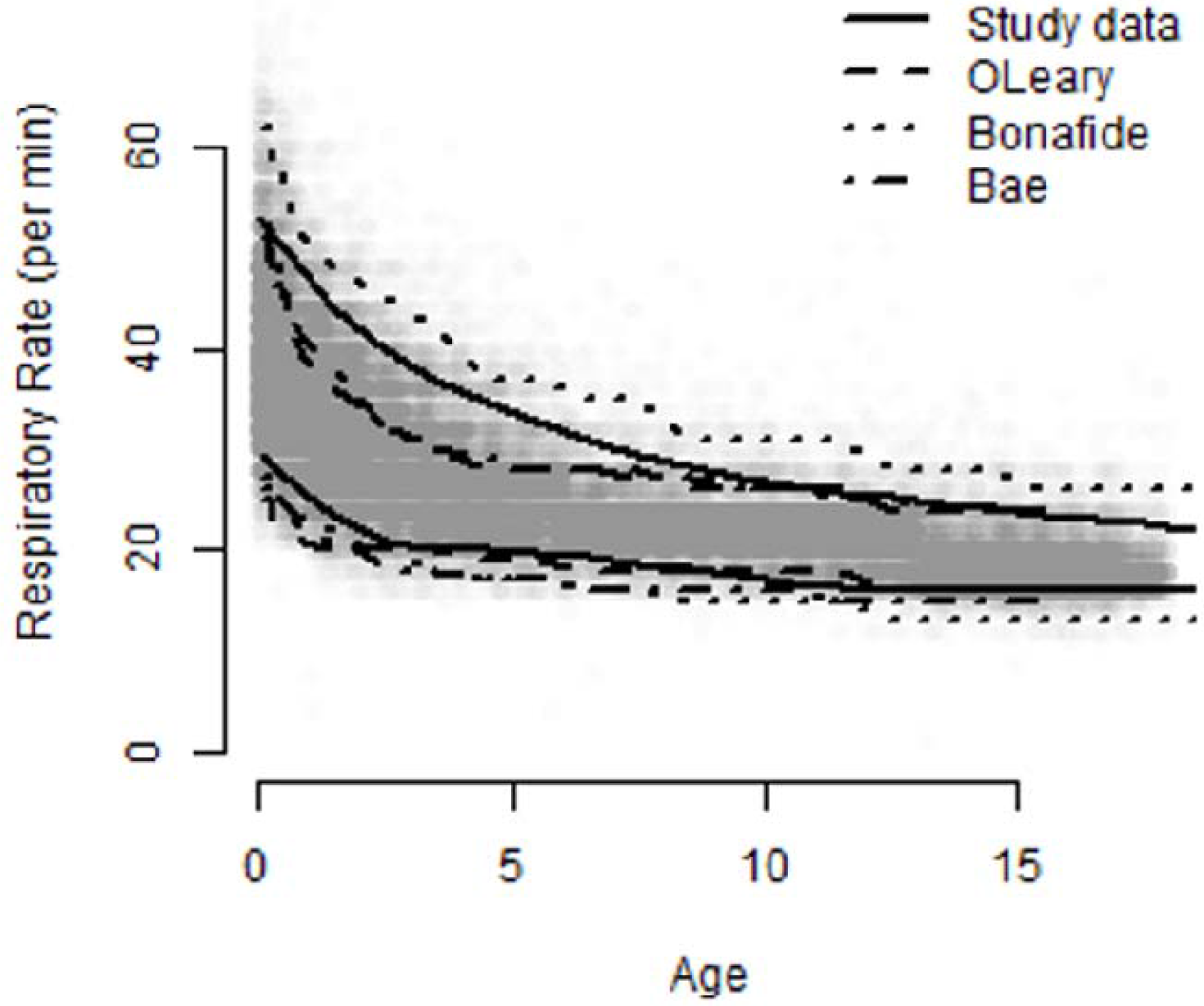
Comparison between the 5^Th^ and 95^th^ centiles of the study cohort data and the 5^th^ and 95^th^ centiles for key papers for respiratory rate.

The study group data is broadly in agreement with APLS ranges for heart rate (Figure 2). There is minor variation between the corresponding 5th and 95th centiles, with the study group’s 50th centile sitting centrally between the two. The study group heart rate sits up to 9 beats /minute higher on the 95th centile in the three months to five years age groups, for all other age groups there is little difference. There are also minimal variations between the 5th centiles. APLS 8 reports a lower and upper band of systolic blood pressure, for simplicity we compare the lower end of the lower band to our 5th centile, and the upper end of the upper band to our 95th centile. The study group has higher SBP than APLS’s lowest reference ranges in all age groups. Our SBP is higher in the younger years, but generally in agreement thereafter, although the upper limit for the over 12-years is higher than our 95th centile.

Sensitivity analyses of (i) RR (n = 33,888) excluding respiratory diagnoses, (ii) HR (n = 39,865) and (iii) SBP (n = 11,593) excluding cardiac diagnoses. The plots of these analyses are presented in Figure 4.

**Figure 4:**
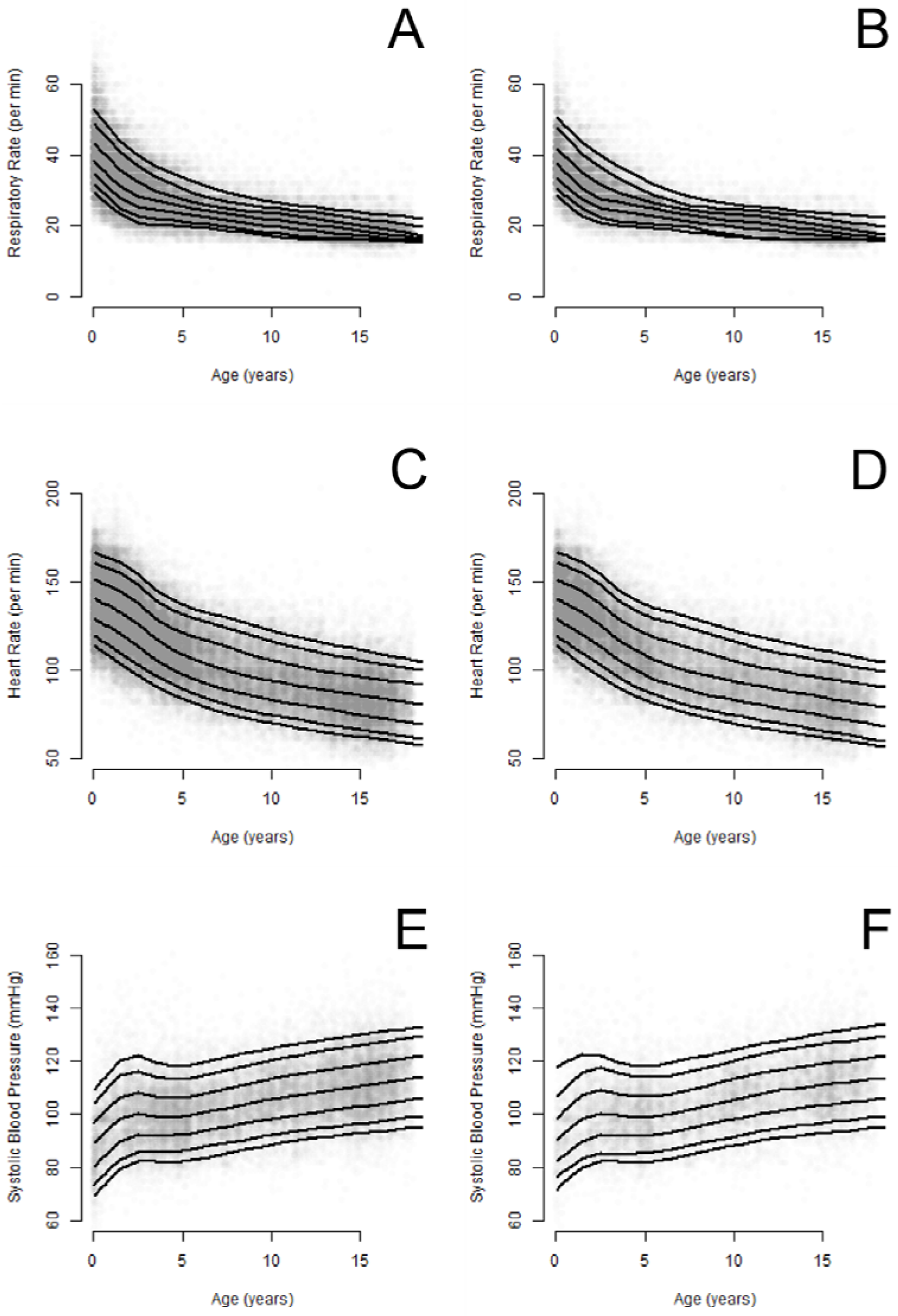
Comparison between main analysis and sensitivity analyses of respiratory rate, heart rate and systolic blood pressure. A, C, and E are the main analysis, and B, D and F are subgroup analyses. For respiratory rate, we present the respiratory rate of children excluding those with respiratory disease (B). For heart rate (D) and systolic blood pressure (F), we excluded children with cardiac disease.

## DISCUSSION

Accurate, contemporary, reference ranges for observations in children are central to the practice of pediatrics. We present data on 66,353 sets of observations conducted at discharge from a busy acute tertiary children’s hospital. Our large dataset allowed us to create robust centile charts of what we believe are ‘safe for discharge’ observations.

We have found a high variability of RR, particularly in the younger age groups. This is in line with respiratory disease being a common cause of paediatric admission.^16^ Our HR data is in keeping with the work of others that as age increases HR decreases in a monotonically inverse rather than linear matter.^1,3,7^ We also note that our SBP centile charts have a peak at two years of age, and after plateau then a rise from six years of age onwards. We did not identify any description of such pattern in the literature.

We identify important differences between our data and the reference ranges of the ALSG,^8^ which are widely used in UK practice. RR and SBP centiles are particularly different, especially among younger age groups. These findings are consistent with those of Chan et al.^17^ who found that approximately 50% of patients had higher SBP compared to APLS ranges. However, Chan et al used a previous version of APLS reference ranges and conducted their study in China.^8^

The 5^th^ centiles are relatively closely aligned and if the step wise APLS ranges were smoothed then this would demonstrate even more alignment between the two centiles. However, the stepped approach used in APLS reference ranges, results in divergence between our centile charts and their reference ranges at the age-based transition points.^8^ This is particularly significant for the younger age groups where variation in physiology and development changes rapidly, this pace of change decreases with age.^18^ We argue that whilst in routine clinical practice having simple age bands with upper and lower limits is a practical and sensible approach to interpreting observations, a reference range approach may result in false-positive and -negative scores at transition points.

As the clinicians taking the observations were unaware of the study, we contend that our data are broadly representative of real-world observations. However, a key limitation of our study is that we used vital sign information at the point of discharge from hospital. This could be an explanation for the high RR of our cohort, as children will likely have not completely recovered from a respiratory illness at the point of discharge (we note that respiratory illness accounts for a substantial proportion of paediatric admissions). However, when Fleming et al.^1^ conducted a meta-analysis using observations from healthy individuals outside of the hospital setting, the 50^th^ centile they found was above ours. It was up to six breaths per minute higher than ours.

We used secondary data which are limited by the availability of data to investigate the associations of co-variables and vital signs. Although measurement errors are a potential study limitation, the hospital aimed to minimize these by having local measures in place to maintain standards of practice regarding the measuring and recording of vital signs. These measures include: the hospital’s adoption of a vital sign policy; the required training of staff; and the standardization of methods and equipment and its ongoing maintenance.

Future research is required to understand the associations between patient characteristics, study setting, and diagnosis to develop understanding and vital sign centiles for specific situations. The increasing availability of substantial amounts of routinely collected data will provide the opportunity to explore trends of vital signs (such as improvement after admission) to identify patients who are not responding to acute therapy, and patients who have deteriorated. This will provide useful additional insights that will further support the development of evidence-based centiles for vital signs, and their utility in clinical practice.

## CONCLUSION

We present centile charts for HR, RR and BP of children and young people at the end of a hospital admission and suggest that comparing acute admission observations to our centile charts could help in the assessment of gauging the severity of paediatric illness. We contrast our centile charts with standard reference ranges from key groups, and from literature and identify significant differences in published reference ranges. The contemporary centiles we derived may serve as useful references for clinicians and could be used to inform the development of evidence-based parameters for vital sign monitor limits. Future work needs to understand the utility of these centiles in different healthcare settings.

## Data Availability

All data produced in the present study are available upon reasonable request to the authors

## Statements and Declarations

### Funding source

This research was funded through a personal award (Taylor) from the National Institute for Health and Care Research, UK.

### Availability of data and material

Anonymized dataset available on request from the corresponding author.

### Code availability

Code for data analysis available on request from the corresponding author.

### Ethics approval

NHS HRA REC approval not required for study. Institutional Governance Approval obtained from Nottingham University Hospitals NHS Trust.

### Consent to participate

N/A

### Consent for publication

N/A

### Contributor statement

Taylor conceptualized and designed the study, managed, and cleaned the dataset, conducted data analyses, drafted the initial manuscript, and reviewed and revised the manuscript. Prayle conducted data analyses, drafted the initial manuscript, and reviewed and revised the manuscript. Wark, Coad and Manning conceptualized and designed the study, supervised data management and analysis, drafted the initial manuscript, and reviewed and revised the manuscript. All authors approved the final manuscript as submitted and agree to be accountable for all aspects of the work.

### Declaration of Interest

Dr Joseph C. Manning is a current recipient of an NIHR (National Institute for Health and Social Care Research) HEE (Health Education England) funded ICA (Integrated Clinical Academic) Clinical Lectureship [ICA-CL-2018-04-ST2-009]. The views expressed in this article are those of the authors and not necessarily those of the NIHR or Department of Health and Social Care, UK. All other authors have no conflicts of interest relevant to this article to disclose.

## Notes

### Author Declarations

NHS Health Research Authority Research Ethics committee approval was not required for this study. Institutional Governance Approval was obtained from Nottingham University Hospitals NHS Trust.

